# Resting Heart Rate, Electrocardiographic Markers of Atrial Cardiopathy, and All-Cause Mortality

**DOI:** 10.64898/2026.03.02.26347372

**Authors:** Paul J. Chu, Mohamed A. Mostafa, Patrick Cheon, Mai Z. Soliman, Elsayed Z. Soliman

## Abstract

**Background:** Elevated resting heart rate (HR) and atrial cardiopathy are each linked to higher mortality risk, yet their interrelationship and joint prognostic value remain unclear.

**Methods:** We analyzed 7,326 adults (mean age 59 ± 13 years) without cardiovascular disease from the Third National Health and Nutrition Examination Survey with available electrocardiograms. Atrial cardiopathy was defined by electrocardiogram as abnormal P-wave axis or deep terminal P-wave negativity in V1. Multivariable logistic regression assessed cross-sectional associations between HR categories and atrial cardiopathy. Cox proportional hazards models evaluated independent and joint associations of HR categories and atrial cardiopathy with all-cause mortality.

**Results:** Atrial cardiopathy was present in 1,833 participants (13.5%). After adjustment, sinus tachycardia (≥100 bpm) was associated with higher odds of atrial cardiopathy (OR 1.76, 95% CI 1.06–2.92), whereas sinus bradycardia (≤50 bpm) was associated with lower odds (OR 0.61, 95% CI 0.43–0.84). Each 10-bpm HR increase corresponded to 25% higher odds of atrial cardiopathy. Over a median 13.8-year follow-up, 2,415 deaths (33.0%) occurred. Sinus tachycardia (HR 3.58, 95% CI 2.61–4.91) and atrial cardiopathy (HR 1.27, 95% CI 1.16–1.39) were independently associated with mortality. Individuals with both conditions had the highest risk (HR 4.11, 95% CI 2.63–6.41). Associations varied by age and race.

**Conclusions:** Elevated resting HR is associated with higher odds of atrial cardiopathy, and their coexistence confers markedly increased mortality risk. Integrating resting HR into atrial cardiopathy metrics may enable granular population-level risk profiling.

## Introduction

Atrial cardiopathy is a clinical construct referring to structural, electrical, or functional abnormalities of the atria that may represent subclinical disease even in the absence of overt atrial fibrillation.^1^,^2^ Elevated resting heart rate (HR) has been closely associated with cardiovascular disease and mortality; however, its relationship with atrial cardiopathy and overall mortality remains incompletely understood.

Although atrial cardiopathy is increasingly recognized as a marker of subclinical disease, simple and readily detectable clinical features that can identify individuals at risk remain poorly defined. Resting HR is a routinely measured vital sign that is widely accessible through wearable devices and affordable home monitoring tools.^4^ It reflects autonomic tone, cardiorespiratory fitness, and subclinical cardiovascular burden, and may therefore serve as an early indicator of cardiovascular health.

We hypothesized that elevated resting HR is associated with increased likelihood of atrial cardiopathy and that both conditions, individually and in combination, are associated with higher mortality risk in the general population. Accordingly, this study aimed to evaluate the relationship between resting HR and atrial cardiopathy and to examine their independent and joint associations with all-cause mortality using data from the National Health and Nutrition Examination Survey (NHANES).

## Methods

### Study population

We used data from participants in the Third National Health and Nutrition Examination Survey (NHANES III), a nationally based survey designed to assess the health and nutritional status of the U.S. civilian population. NHANES III was conducted between 1988 and 1994 by the National Center for Health Statistics (NCHS), Centers for Disease Control and Prevention. All participants provided written informed consent, and the study protocol was approved by the NCHS Institutional Review Board. Detailed information regarding survey design, methodology, and data availability has been described previously.^5^,^6^

For the present analysis, we included participants with available baseline electrocardiograms (ECGs). By design, only individuals aged ≥40 years underwent ECG recording in NHANES III. We excluded participants with prior cardiovascular disease (myocardial infarction, heart failure, or stroke), non-sinus rhythm, or use of antiarrhythmic medications. To minimize misclassification of atrial cardiopathy markers, we additionally excluded participants with major ECG abnormalities defined by Minnesota codes, including atrial fibrillation, atrial flutter, paced rhythm, complete left or right bundle branch block, or significant conduction delays that could distort P-wave morphology. After exclusions, 7,326 participants were included.

### ECG parameters

Standard 12-lead ECGs were recorded during mobile examination center visits using a Marquette MAC-12 electrocardiograph (Marquette Medical Systems, Milwaukee, WI, USA). Digital ECG files were transmitted to the Epidemiological Cardiology Research Center (EPICARE), Wake Forest University School of Medicine, for centralized processing. Each ECG underwent visual inspection by trained technicians followed by automated analysis using the GE 12-SL 2001 program.

Consistent with prior studies, atrial cardiopathy was defined by the presence of either (1) abnormal P-wave axis outside 0°–75°, or (2) deep terminal negativity of the P wave in lead V1 (DTNPV1) exceeding −100 µV on resting 12-lead ECG.^3^,^7^,^8^ The terminal negative amplitude of the P wave in V1 was obtained from automated measurements; values more negative than −100 µV were considered abnormal. Prior studies have demonstrated excellent agreement between automated and manual DTNPV1 measurements.^9^

Resting HR was derived from baseline ECGs and expressed as beats per minute (bpm). Participants were categorized as normal HR (>50–<100 bpm), sinus bradycardia (≤50 bpm), or sinus tachycardia (≥100 bpm).

### Ascertainment of all-cause mortality

The primary outcome was all-cause mortality, determined through linkage to the National Death Index with follow-up through December 31, 2015.

### Other variables

Demographic characteristics (age, sex, race/ethnicity), smoking status, education, family income, and medication use were self-reported during home interviews. Body mass index (BMI) was calculated as weight in kilograms divided by height in meters squared from standardized physical examinations. Blood pressure was measured in the seated position, and the average of up to three readings was used.

Diabetes mellitus was defined as fasting plasma glucose ≥126 mg/dL or use of glucose-lowering medication. Hypertension was defined as systolic BP ≥130 mmHg, diastolic BP ≥80 mmHg, or use of antihypertensive medication. Dyslipidemia was defined as total cholesterol ≥200 mg/dL, triglycerides ≥150 mg/dL, or use of lipid-lowering therapy. Thyroid disease was defined as thyroid-stimulating hormone ≤0.45 mIU/L with free thyroxine >1.6 ng/dL. Cancer history was based on self-reported physician diagnosis of hematologic or solid organ malignancy. Laboratory measurements, including total cholesterol, HDL cholesterol, serum creatinine, and glucose, were obtained using standardized NCHS procedures.^6^

### Statistical analysis

Participant characteristics were compared by atrial cardiopathy status using the chi-square test for categorical variables and Student’s t-test for continuous variables. Categorical variables are presented as counts and percentages, and continuous variables as mean ± standard deviation or median (interquartile range), as appropriate.

Multivariable logistic regression assessed cross-sectional associations between HR and atrial cardiopathy. HR was modeled both categorically (normal, bradycardia, tachycardia) and continuously (per 10-bpm increase). Model 1 adjusted for sociodemographic factors (age, sex, race/ethnicity, education). Model 2 additionally adjusted for smoking status, total cholesterol, lipid-lowering medication use, antihypertensive medication use, systolic BP, BMI, serum creatinine, diabetes mellitus, and thyroid disease. Similar models evaluated associations between HR and individual atrial cardiopathy components (P-wave axis and DTNPV1).

Multivariable Cox proportional hazards models estimated associations of HR categories and atrial cardiopathy, individually and jointly, with all-cause mortality. Covariate adjustment followed the same structure as logistic models; Model 2 additionally included cancer history.

Analyses were performed without NHANES sampling weights; therefore, results reflect the analytic cohort and are not nationally representative. Statistical analyses were conducted using jamovi (Version 2.4.14), and figures were generated using R version 4.5.1. Two-sided P <0.05 denoted statistical significance.

## Results

Our study included 7,326 participants (mean age 59.0 ± 13.3 years; 53.2% women; 48.8% non-Hispanic White). At baseline, atrial cardiopathy was present in 1,833 participants (13.5%). Compared with those without atrial cardiopathy, affected participants were older, more likely to be current smokers, and more likely to have thyroid disease. They also had higher resting heart rate and a greater prevalence of sinus tachycardia. No significant differences were observed in sex distribution. Additional baseline characteristics are shown in **Table 1**.

**Table 1:** population characteristics.

| **Table1: population characteristics** |  |  |  |  |
| --- | --- | --- | --- | --- |
| **Characteristics:**<br>Numbers (%) or<br>Mean (SD) / Median (IQR) | Total<br>(n=7326) | Atrial<br>Cardiopathy<br>Absent<br>(n=5493) | Atrial<br>Cardiopathy<br>Present<br>(n=1833) | P value |
| Age, years | 59 (13.3) | 58.2 (13.2) | 61.7 (13.5) | <0.001 |
| Female | 3900 (53.2) | 2909 (52.96) | 991 (54.06) | 0.493 |
| Race-Ethnicity: |  |  |  |  |
| Non-Hispanic White | 3577 (48.8) | 2579 (46.95) | 998 (54.45) | <0.001 |
| Non-Hispanic Black | 1731 (23.6) | 1241 (22.59) | 490 (26.73) | <0.001 |
| Mexican-American | 1720 (23.5) | 1449 (26.38) | 271 (14.78) | <0.001 |
| Other (%) | 298 (4.1) | 224 (4.08) | 74 (4.04) | 0.895 |
| Education years | 10.4 (4.37) | 10.4 (4.42) | 10.6 (4.21) | 0.047 |
| Family income <20k | 3308 (45.2) | 2443 (44.47) | 865 (47.19) | 0.058 |
| Current Smoker | 1681 (22.9) | 1090 (19.84) | 591 (32.24) | <0.001 |
| Past Smoker | 2301 (31.4) | 1754 (31.93) | 547 (29.84) | 0.138 |
| Use of lipid lowering | 219 (3) | 170 (3.09) | 49 (2.67) | 0.340 |
| Use of BP Medications | 1619 (22.1) | 1263 (22.99) | 356 (19.42) | 0.002 |
| Body Mass Index (kg/m<sup>2</sup>) | 27.6 (5.61) | 28.4 (5.49) | 25.2 (5.22) | <0.001 |
| Serum Creatinine, mg/dl | 1.06(0.485) | 1.06 (0.48) | 1.05 (0.499) | 0.526 |
| Systolic Blood Pressure (mm Hg) | 132 (19.9) | 132 (19.6) | 132 (20.6) | 0.568 |
| Diastolic Blood Pressure (mm Hg) | 76.4 (10.4) | 77 (10.4) | 74.8 (10.1) | <0.001 |
| Triglycerides (mg/dL) | 125 (57.1) | 130 (47) | 110 (71) | <0.001 |
| Total Cholesterol (mg/dL) | 209 (59.3) | 211 (58.5) | 206 (61.9) | 0.005 |
| HDL Cholesterol (mg/dL) | 48.9 (19.2) | 47.7 (18.3) | 52.6 (21.3) | <0.001 |
| HR, BPM | 68.6 (11.6) | 68 (11.3) | 70.2 (12.3) | <0.001 |
| P-wave axis degree | 58.7 (22.6) | 53.3 (16.9) | 75 (28.8) | <0.001 |
| Cancer history | 748 (10.2) | 534 (9.7) | 214 (11.6) | 0.025 |
| Thyroid disease | 381 (5.2) | 262 (4.77) | 119 (6.49) | 0.004 |
| Diabetes Mellitus | 787 (10.7) | 633 (11.52) | 154 (8.40) | <0.001 |
| Hypertension | 2508 (34.2) | 1966 (35.79) | 542 (29.57) | <0.001 |
| Dyslipidemia | 1699 (23.2) | 1313 (23.90) | 386 (21.06) | 0.010 |
| Heart Rate Groups: |  |  |  |  |
| Normal Heart Rate | 7005 (95.6) | 5251 (95.59) | 1754 (95.69) | <0.001 |
| Sinus Bradycardia | 246 (3.4) | 196 (3.57) | 50 (2.73) | <0.001 |
| Sinus Tachycardia | 75 (1) | 46 (0.84) | 29 (1.58) | <0.001 |
| Bradycardia: ≤ 50 BPM<br>Normal Heart Rate: >50 - <100 BPM<br>Tachycardia: ≥ 100 BPM<br>Data presented as mean (SD) for normally distributed continuous data or median (IQR) for skewed data. Categorical data presented as numbers and percentage. |  |  |  |  |

In multivariable logistic regression adjusting for sociodemographic and cardiovascular risk factors, sinus tachycardia was independently associated with higher odds of atrial cardiopathy (OR 1.76, 95% CI 1.06–2.92), whereas sinus bradycardia was inversely associated (OR 0.61, 95% CI 0.43–0.84). When modeled continuously, each 10-bpm increase in heart rate was associated with a 25% higher odds of atrial cardiopathy (**Table 2**). Analyses of individual atrial cardiopathy components showed similar patterns for abnormal P-wave axis, with positive associations for tachycardia and inverse associations for bradycardia, whereas heart-rate categories were not significantly associated with DTNPV1. These component-level analyses are presented in **Supplementary Tables 1 and 2**.

**Table 2:**
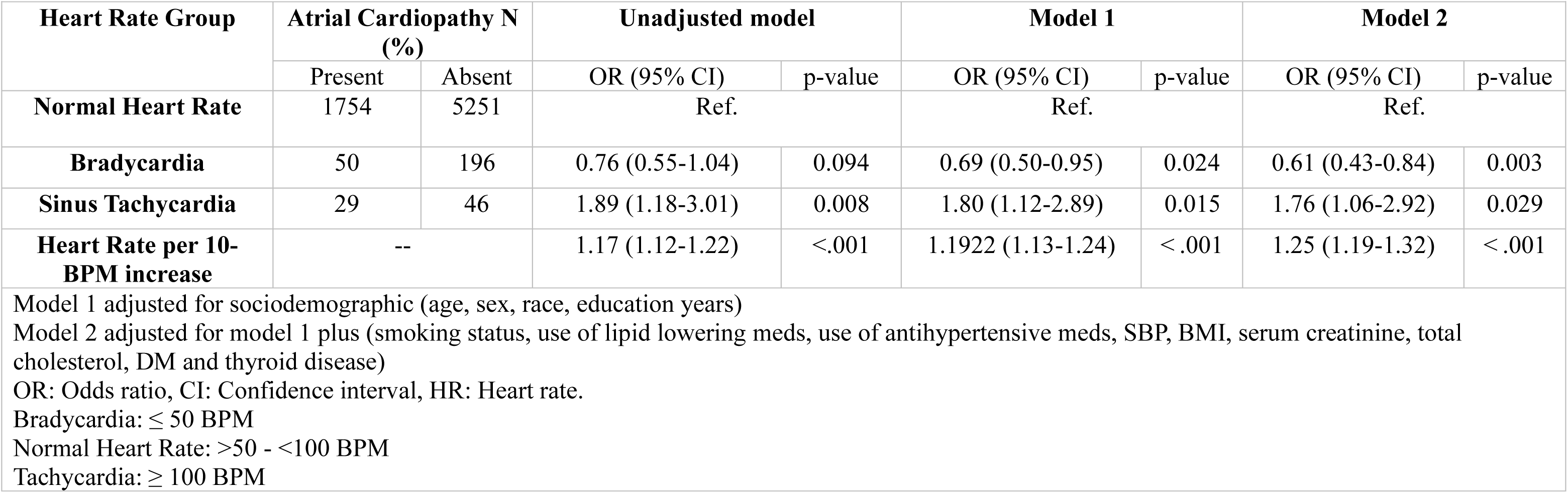
Association between Resting Heart Rate Groups and Atrial Cardiopathy.

| Heart Rate Group | Atrial Cardiopathy N (%) |  | Unadjusted model |  | Model 1 |  | Model 2 |  |
| --- | --- | --- | --- | --- | --- | --- | --- | --- |
|  | Present | Absent | OR (95% CI) | p-value | OR (95% CI) | p-value | OR (95% CI) | p-value |
| **Normal Heart Rate** | 1754 | 5251 | Ref. |  | Ref. |  | Ref. |  |
| **Bradycardia** | 50 | 196 | 0.76 (0.55-1.04) | 0.094 | 0.69 (0.50-0.95) | 0.024 | 0.61 (0.43-0.84) | 0.003 |
| **Sinus Tachycardia** | 29 | 46 | 1.89 (1.18-3.01) | 0.008 | 1.80 (1.12-2.89) | 0.015 | 1.76 (1.06-2.92) | 0.029 |
| **Heart Rate per 10-BPM increase** | -- |  | 1.17 (1.12-1.22) | <.001 | 1.1922 (1.13-1.24) | <.001 | 1.25 (1.19-1.32) | <.001 |

Model 1 adjusted for sociodemographic (age, sex, race, education years)
Model 2 adjusted for model 1 plus (smoking status, use of lipid lowering meds, use of antihypertensive meds, SBP, BMI, serum creatinine, total cholesterol, DM and thyroid disease)
OR: Odds ratio, CI: Confidence interval, HR: Heart rate.
Bradycardia: $\leq 50$ BPM
Normal Heart Rate: $>50 - <100$ BPM
Tachycardia: $\geq 100$ BPM

During a median follow-up of 13.8 years, 2,415 deaths (33.0%) occurred. In fully adjusted Cox models, sinus tachycardia was associated with increased all-cause mortality compared with normal heart rate (HR 3.58, 95% CI 2.61–4.91), whereas bradycardia was not significantly associated with mortality. Each 10-bpm increase in heart rate corresponded to approximately 19% higher mortality risk (**Table 3**). Atrial cardiopathy was also independently associated with increased mortality risk (HR 1.27, 95% CI 1.16–1.39) (**Table 4**).

**Table 3:** Association Between Resting Heart Rate and All-Cause Mortality.

| Exposure: | Participants/<br>events | Crude Model |  | Model 1 |  | Model 2 |  |
| --- | --- | --- | --- | --- | --- | --- | --- |
|  |  | HR (95% CI) | p-value | HR (95% CI) | p-value | HR (95% CI) | p-value |
| **Normal Heart Rate** | 7005/2281 | Reference | -- | Reference | -- | Reference | -- |
| **Bradycardia** | 246/86 | 1.06 (0.85-1.31) | p=0.619 | 0.81 (0.65-1.01) | 0.057 | 0.83 (0.66-1.03) | 0.091 |
| **Sinus Tachycardia** | 75/48 | 2.77 (2.03-3.79) | <0.001 | 4.04 (2.95-5.54) | <0.001 | 3.58 (2.61-4.91) | <0.001 |
| **Heart Rate per 10-BPM increase** | -- | 1.12 (1.08-1.16) | <0.001 | 1.20 (1.16-1.24) | <0.001 | 1.19 (1.15-1.23) | <0.001 |

Model 1 adjusted for sociodemographic (age, sex, race, education years)
Model 2 adjusted for model 1 plus (smoking status, use of lipid lowering meds, use of antihypertensive meds, SBP, BMI, serum creatinine, total cholesterol, DM, thyroid disease and cancer)
HR: Hazard ratio, CI: Confidence interval, HR: Heart rate.
Bradycardia: $\leq 50$ BPM
Normal Heart Rate: $>50 - <100$ BPM
Tachycardia: $\geq 100$ BPM

**Table 4:** Association Between Atrial Cardiopathy and All-cause Mortality.

| Exposure | Participants/events | Crude Model |  | Model 1 |  | Model 2 |  |
| --- | --- | --- | --- | --- | --- | --- | --- |
|  |  | HR (95% CI) | p-value | HR (95% CI) | p-value | HR (95% CI) | p-value |
| **Atrial Cardiopathy Absent** | 5493/1626 | Reference | -- | Reference | -- | Reference | -- |
| **Atrial Cardiopathy Present** | 1833/789 | 1.66 (1.53-1.81) | <0.001 | 1.35 (1.24-1.47) | <0.001 | 1.27 (1.16-1.39) | <0.001 |

Model 1 adjusted for sociodemographic (age, sex, race, education years) Model 2 adjusted for model 1 plus (smoking status, use of lipid lowering meds, use of antihypertensive meds, SBP, BMI, serum creatinine, total cholesterol, DM, thyroid disease and cancer) HR: Hazard ratio, CI: Confidence interval

Joint analyses demonstrated a graded increase in mortality risk across combinations of heart-rate categories and atrial cardiopathy. Participants with both sinus tachycardia and atrial cardiopathy had the highest mortality risk compared with those with normal heart rate and no atrial cardiopathy (HR 4.11, 95% CI 2.63–6.41). Tachycardia without atrial cardiopathy and normal heart rate with atrial cardiopathy were also associated with elevated risk, though of lesser magnitude (**Table 5**). Adjusted survival curves illustrating these differences are presented in **Figure 1**.

**Table 5:** Association of Resting Heart Rate Groups and Atrial Cardiopathy Combinations and All-Cause Mortality.

| **Exposure** | **Participants/**<br>**events** | **Crude Model**<br>**Hazard Ratio (95% CI)** | **Model 1**<br>**Hazard Ratio (95% CI)** | **Model 2**<br>**Hazard Ratio (95% CI)** |
| --- | --- | --- | --- | --- |
| **Normal HR + Absent Atrial cardiopathy** | 5251/1530 | Reference | Reference | Reference |
| **Bradycardia + Absent Atrial cardiopathy** | 196/68 | 1.18 (0.92-1.50, p=0.186) | 0.83 (0.65-1.06, p=0.141) | 0.82 (0.64-1.05, p=0.109) |
| **Bradycardia + Present Atrial cardiopathy** | 50/18 | 1.34 (0.84-2.13, p=0.219) | 1.16 (0.73-1.84, p=0.539) | 1.09 (0.68-1.74, p=0.716) |
| **Normal HR + Present Atrial cardiopathy** | 1754/751 | 1.63 (1.49-1.78, p<0.001) | 1.31 (1.20-1.43, p<0.001) | 1.24 (1.13-1.36, p<0.001) |
| **Tachycardia + Absent Atrial cardiopathy** | 46/28 | 3.12 (2.14-4.53, p<0.001) | 3.72 (2.56-5.41, p<0.001) | 3.49 (2.40-5.08, p<0.001) |
| **Tachycardia + Present Atrial cardiopathy** | 29/20 | 3.74 (2.41-5.82, p<0.001) | 5.20 (3.34-8.09, p<0.001) | 4.11 (2.63-6.41, p<0.001) |

HR: heart rate, CI: Confidence interval
Model 1 adjusted for sociodemographic (age, sex, race, education years)
Model 2 adjusted for model 1 plus (smoking status, use of lipid lowering meds, use of antihypertensive meds, SBP, BMI, serum creatinine, total cholesterol, DM, thyroid disease and Cancer)
Bradycardia: $\leq 50$ BPM
Normal Heart Rate: $>50 - <100$ BPM
Tachycardia: $\geq 100$ BPM

Subgroup analyses revealed significant effect modification by age and race, but not by sex, with stronger associations between heart rate and atrial cardiopathy observed among participants <65 years and among Black participants (**Table 6**).

**Table 6:** Subgroup Analysis for the Association between Heart rate and Atrial Cardiopathy.

|  |  |  |  |
| --- | --- | --- | --- |
| Subgroup: |  | Odds ratio per 10-bpm increase (95% CI) * ‡ | p-interaction |
| Age | <65 years | 1.34 (1.26–1.44) | 0.001 |
|  | ≥65 years | 1.15 (1.07–1.23) |  |
| Sex | Men | 1.27 (1.19–1.36) | 0.79 |
|  | Women | 1.25 (1.14–1.37) |  |
| Race | Non-Black | 1.21 (1.14–1.28) | 0.008 |
|  | Black | 1.41 (1.25–1.59) |  |
| OR: Odds ratio. CI: confidence interval. bpm: beat per minute |  |  |  |
| *Odds ratios reported per 10 BPM increase in heart rate |  |  |  |
| ‡Model adjusted for age, sex, race, education years, smoking status, use of lipid lowering meds, use of antihypertensive meds, SBP, BMI, serum creatinine, total cholesterol, DM, and thyroid disease. |  |  |  |

## Discussion

In this cohort of U.S. adults without baseline cardiovascular disease, resting heart rate and atrial cardiopathy demonstrated a strong cross-sectional association, with higher heart rate linked to greater odds of atrial cardiopathy and lower heart rate associated with reduced odds. Longitudinally, both elevated resting heart rate and atrial cardiopathy were independently associated with all-cause mortality, and their coexistence conferred the highest risk. These findings suggest that atrial cardiopathy may represent a shared biological substrate through which elevated heart rate and adverse outcomes intersect, although temporal directionality cannot be established from the present analysis.

In prior work, elevated resting heart rate and electrocardiographic markers of atrial cardiopathy were each independently associated with incident atrial fibrillation. Because atrial fibrillation is itself a well-established predictor of mortality, evaluating upstream atrial substrates and physiologic signals is clinically relevant. The present study extends this line of investigation by examining the interrelationship between resting heart rate and atrial cardiopathy and by evaluating their joint association with all-cause mortality in individuals without overt atrial fibrillation.

Our findings indicate that resting heart rate is associated with multiple electrocardiographic features of atrial cardiopathy, particularly abnormal P-wave axis, even in the absence of clinical arrhythmia. Several biological mechanisms may underlie these relationships. Elevated heart rate shortens diastolic filling time, increases sympathetic activation, and may promote profibrotic signaling, all of which contribute to atrial structural and electrical remodeling. Experimental pacing models have demonstrated that sustained atrial tachycardia induces electrical remodeling through downregulation of L-type calcium currents and shortening of the atrial effective refractory period, leading to conduction heterogeneity and delayed atrial activation. These processes are detectable on surface electrocardiography as altered P-wave morphology before the onset of atrial fibrillation. Conversely, lower resting heart rate may reflect reduced atrial rate stress and preservation of normal refractory properties, potentially supporting structural and electrical integrity. Prospective studies incorporating repeated heart-rate and ECG measurements are needed to clarify temporal sequence and cumulative rate exposure.

Consistent with prior population-based investigations, elevated resting heart rate was strongly associated with all-cause mortality in a graded fashion. Resting heart rate reflects the integrated effects of autonomic balance, cardiorespiratory fitness, and subclinical cardiovascular burden. Mechanistic studies have linked chronic sympathetic predominance and reduced parasympathetic tone to oxidative stress, endothelial dysfunction, and accelerated atherosclerosis, pathways plausibly contributing to excess mortality. These observations reinforce resting heart rate as a clinically meaningful physiologic marker rather than a benign vital sign.

Atrial cardiopathy also demonstrated an independent association with mortality after adjustment for demographic and clinical risk factors. Previous studies have primarily linked atrial cardiopathy to stroke and thromboembolic outcomes; the present findings broaden this perspective by demonstrating associations with all-cause mortality, suggesting that atrial cardiopathy reflects a wider burden of subclinical cardiovascular disease. Structural and electrical atrial remodeling promotes chamber dysfunction, blood stasis, and a prothrombotic milieu while simultaneously serving as a marker of systemic vascular and metabolic stress. Thus, atrial cardiopathy likely represents both a localized atrial process and a manifestation of generalized cardiovascular vulnerability.

Joint analyses provided additional insight, demonstrating a stepwise increase in mortality risk across combinations of heart-rate categories and atrial cardiopathy, with the greatest risk observed when both conditions coexisted. This pattern is consistent with overlapping and potentially synergistic pathophysiologic pathways. Elevated heart rate may function both as a marker of autonomic imbalance and as a contributor to atrial remodeling, whereas atrial cardiopathy reflects structural substrate vulnerability. Their coexistence may therefore identify a high-risk phenotype characterized by heightened electrical, structural, and systemic stress.

We observed significant effect modification by age and race but not by sex. The association between heart rate and atrial cardiopathy was stronger in adults younger than 65 years, possibly reflecting earlier detection of rate-related remodeling before age-related structural disease predominates. Stronger associations among Black participants align with prior observations of racial heterogeneity in atrial structural and electrical phenotypes, potentially related to differences in fibrosis burden, autonomic tone, and comorbidity patterns. These findings underscore the importance of demographic context when interpreting heart-rate–related risk signals.

From a clinical perspective, resting heart rate is measured routinely and requires no additional resources. In this study, sinus tachycardia frequently co-occurred with electrocardiographic markers of atrial cardiopathy that are readily identifiable on standard 12-lead ECGs but often overlooked. Integrating heart rate with targeted P-wave assessment may represent a low-cost strategy to identify individuals with subclinical atrial disease and elevated mortality vulnerability. While these findings do not establish that modifying heart rate will alter outcomes, they support greater attention to resting heart rate as part of cardiovascular risk evaluation.

Several questions remain. First, the cross-sectional assessment of heart rate and atrial cardiopathy precludes determination of temporal sequence. Longitudinal studies with repeated ECG and physiologic measurements are needed. Second, whether pharmacologic heart-rate reduction can prevent or reverse ECG-defined atrial cardiopathy in the general population is unknown. Third, ECG-based definitions capture only part of atrial disease; future work incorporating cardiac magnetic resonance imaging, echocardiographic atrial strain, and circulating biomarkers would provide a more comprehensive assessment. Finally, evaluation of cause-specific mortality may clarify the biological pathways linking tachycardia, atrial remodeling, and adverse outcomes.

Several limitations warrant consideration. Resting heart rate and atrial cardiopathy were each assessed using single baseline ECG measurements, which may not reflect long-term variability and could introduce misclassification. Residual confounding is possible despite multivariable adjustment. The ECG-based definition of atrial cardiopathy does not capture structural abnormalities detectable by imaging, and findings from this historical cohort may not fully generalize to contemporary or non-U.S. populations. Nonetheless, strengths include the large sample size, standardized ECG acquisition with centralized interpretation, and long-term mortality follow-up.

## Conclusions

In this community-based cohort, both elevated resting heart rate and atrial cardiopathy were independently associated with increased all-cause mortality, with the highest risk observed when both conditions coexisted. These findings highlight resting heart rate as a widely accessible physiologic signal that, when considered alongside electrocardiographic indicators of atrial cardiopathy, may help identify individuals with subclinical atrial disease and elevated mortality risk in the general population.

## Data Availability Statement

Data used in this study are publicly available at https://wwwn.cdc.gov/nchs/nhanes/nhanes3/datafiles.aspx

## Author Approval

All authors have seen and approved the manuscript.

