## Supplemental Table 1 and Supplemental Table 2 for "Resting Heart Rate, Electrocardiographic Markers of Atrial Cardiopathy, and All-Cause Mortality"

| Supp. Table 1: Association between Heart Rate Groups and DTNPV1 | | | | | | | | |
| --- | --- | --- | --- | --- | --- | --- | --- | --- |
| Heart Rate Groups | **PTFV1** | | **Unadjusted model** | | **Model 1** | | **Model 2** | |
|  | **Present** | **Absent** | **OR (95% CI)** | **p-value** | **OR (95% CI)** | **p-value** | **OR (95% CI)** | **p-value** |
| Normal Heart Rate | 172 | 6833 | Ref. | | Ref. | | Ref. | |
| Bradycardia | 3 | 243 | 0.49 (0.15-1.54) | 0.224 | 0.43 (0.13- 1.37) | 0.156 | 0.43 (0.13-1.34) | 0.145 |
| Sinus Tachycardia | 4 | 71 | 2.23 (0.80-6.19) | 0.121 | 2.12 (0.76-5.93) | 0.151 | 2.07 (0.73-5.84) | 0.166 |
| Heart Rate per 10-BPM increase | -- | | 1.19 (1.06-1.34) | 0.002 | 1.20 (1.074-1.35) | < .001 | 1.21 (1.08-1.35) | < .001 |
| DTNPV1: deep terminal negativity of P wave in V1.  Model 1 adjusted for sociodemographic (age, sex, race, education years)  Model 2 adjusted for model 1 plus (smoking status, use of lipid lowering meds, use of antihypertensive meds, SBP, BMI, serum creatinine, total cholesterol, DM, thyroid disease)  OR: Odds ratio, CI: Confidence interval, HR: Heart rate.  Bradycardia: ≤ 50 BPM  Normal Heart Rate: >50 - <100 BPM  Tachycardia: ≥ 100 BPM | | | | | | | | |

| Supp. Table 2: Association between Heart Rate Groups and P Wave Axis | | | | | | | | |
| --- | --- | --- | --- | --- | --- | --- | --- | --- |
| Heart Rate Groups: | **Abnormal P wave Axis** | | **Unadjusted model** | | **Model 1** | | **Model 2** | |
|  | **Present** | **Absent** | **OR (95% CI)** | **p-value** | **OR (95% CI)** | **p-value** | **OR (95% CI)** | **p-value** |
| Normal Heart Rate | 1649 | 5356 | Ref. | | Ref. | | Ref. | |
| Bradycardia | 48 | 198 | 0.78 (0.57-1.08) | 0.143 | 0.71 (0.52-0.99) | 0.046 | 0.63 (0.45-0.88) | 0.007 |
| Sinus Tachycardia | 27 | 48 | 1.83 (1.14-2.94) | 0.013 | 1.74 (1.08-2.82) | 0.023 | 1.70 (1.01-2.86) | 0.043 |
| Heart Rate per 10-BPM increase | -- | | 1.16 (1.117-1.22) | < .001 | 1.18 (1.13-1.24) | < .001 | 1.25 (1.19-1.31) | < .001 |
| Model 1 adjusted for sociodemographic (age, sex, race, education years)  Model 2 adjusted for model 1 plus (smoking status, use of lipid lowering meds, use of antihypertensive meds, SBP, BMI, serum creatinine, total cholesterol, DM, thyroid disease)  OR: Odds ratio, CI: Confidence interval, HR: Heart rate.  Bradycardia: ≤ 50 BPM  Normal Heart Rate: >50 - <100 BPM  Tachycardia: ≥ 100 | | | | | | | | |
